# Artificial intelligence to facilitate clinical trial recruitment in age-related macular degeneration

**DOI:** 10.1101/2024.02.15.24302891

**Authors:** Dominic J. Williamson, Robbert R. Struyven, Fares Antaki, Mark A. Chia, Siegfried K. Wagner, Mahima Jhingan, Zhichao Wu, Robyn Guymer, Simon S. Skene, Naaman Tammuz, Blaise Thomson, Reena Chopra, Pearse A. Keane

## Abstract

**Background:** Recent developments in artificial intelligence (AI) have positioned it to transform several stages of the clinical trial process. In this study, we explore the role of AI in clinical trial recruitment of individuals with geographic atrophy (GA), an advanced stage of age-related macular degeneration, amidst numerous ongoing clinical trials for this condition.

**Methods:** Using a diverse retrospective dataset from Moorfields Eye Hospital (London, United Kingdom) between 2008 and 2023 (602,826 eyes from 306,651 patients), we deployed a deep learning system trained on optical coherence tomography (OCT) scans to generate segmentations of the retinal tissue. AI outputs were used to identify a shortlist of patients with the highest likelihood of being eligible for GA clinical trials, and were compared to patients identified using a keyword-based electronic health record (EHR) search. A clinical validation with fundus autofluorescence (FAF) images was performed to calculate the positive predictive value (PPV) of this approach, by comparing AI predictions to expert assessments.

**Results:** The AI system shortlisted a larger number of eligible patients with greater precision (1,139, PPV: 63%; 95% CI: 54–71%) compared to the EHR search (693, PPV: 40%; 95% CI: 39– 42%). A combined AI-EHR approach identified 604 eligible patients with a PPV of 86% (95% CI: 79–92%). Intraclass correlation of GA area segmented on FAF versus AI-segmented area on OCT was 0.77 (95% CI: 0.68–0.84) for cases meeting trial criteria. The AI also adjusts to the distinct imaging criteria from several clinical trials, generating tailored shortlists ranging from 438 to 1,817 patients.

**Conclusions:** We demonstrate the potential for AI in facilitating automated pre-screening for clinical trials in GA, enabling site feasibility assessments, data-driven protocol design, and cost reduction. Once treatments are available, similar AI systems could also be used to identify individuals who may benefit from treatment.

## Introduction

Bringing a new drug to market is costly and time-consuming. The process from drug discovery to approval can take 10 to 15 years and can cost ∼$1 billion.^1,2^ Clinical trials play an important role in this process, with significant resources dedicated to proving the safety and efficacy of any prospective treatment. Today, challenges in recruiting participants stand as a significant obstacle in clinical trials.^3,4^ Time taken to fully recruit a trial adds significantly to the cost and can lead to delays, with up to 86% of trials not finishing on schedule.^5,6^ Several methods have been proposed to alleviate the recruitment problem, including open-trial designs and the utilization of technology.^4,7^

Age-related macular degeneration (AMD) is a leading cause of vision loss in individuals over 50 years old in developed countries.^8^ One late form of the disease is geographic atrophy (GA) where central vision is lost as a result of cell death. Eventually, GA leads to irreversible vision loss and legal blindness. The disease affects 5 to 10 million people worldwide, with its prevalence projected to increase.^9,10^ Until recently, there were no proven treatments for GA, however two treatments have recently been approved by the U.S. Food and Drug Administration.^11,12^ These treatments show modest decline in the rate of GA area growth, without benefit for visual acuity.^13,14^ As such, many trials of other interventions are underway with others planned to commence in the near future.^15,16^

In clinical trials for GA, a key challenge lies in identifying patients who meet inclusion criteria, which mandates fundus autofluorescence (FAF) imaging – a modality which, in contrast to optical coherence tomography (OCT), is not routinely obtained for AMD patients.^8,17^ Any ability to facilitate these trials, specifically around recruitment times, would help identify efficacious treatments for this devastating disease. Traditional patient recruitment through electronic health records (EHR) searches in clinical notes is often inefficient, requiring significant manual effort to identify patients with GA before validating their diagnosis with FAF imaging. EHRs may also have inaccuracies and inconsistencies, especially in unstructured data, and GA may frequently be underreported. When GA is noted, the records might lack sufficient imaging-based details for meeting strict trial inclusion criteria.^18,19^

Deploying artificial intelligence (AI) methodologies could contribute to the solution.^20^ AI could be used to analyze large EHR datasets to identify potential participants with a high likelihood of eligibility.^7,21^ It could be particularly effective in clinical trials with imaging-based selection criteria, such as in GA, with its ability to extract valuable information from imaging routinely collected during clinical practice. Using AI to help assess retinal images could be a key step towards a more efficient approach to screening and, if successful, this could significantly shorten recruitment times in on-going and future trials.

In this study, we explore the use of AI in identifying patients with GA to facilitate clinical trial recruitment, as described in **Figure 1**. As a primary objective, we compare the outcomes of an AI-based approach with a conventional EHR search in identifying patients with imaging features required for inclusion into specific clinical trials. As a secondary objective, we study the agreement between human-graded GA lesion size on FAF compared to AI segmentation on OCT, and the ability to filter for additional selection criteria.

**Figure 1.**
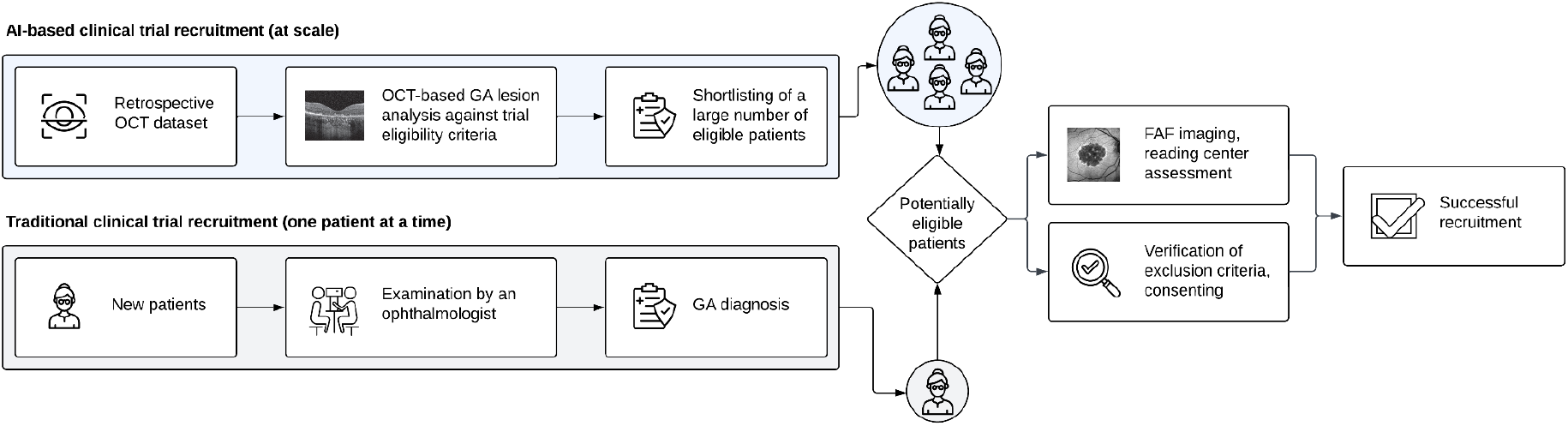
Potential use case of AI for clinical trial recruitment in the setting of geographic atrophy. Compared to traditional methods of recruitment that allow the identification of a single patient at a time, AI can sift through large retrospective imaging datasets and shortlist patients using trial-specific imaging criteria.

## Methods

### Dataset and participants

We used a large retrospective dataset from the INSIGHT Health Data Research Hub at Moorfields Eye Hospital in London, United Kingdom. The dataset comprised 306,651 patients (602,826 eyes) who attended Moorfields between 1 January 2008 until 10 April 2023 with suspected retinal disease and underwent OCT imaging as part of their clinical assessment. A total of 78,917 patients (154,410 eyes) remained after applying the inclusion criteria detailed in the **Supplementary Appendix. Figure S1** illustrates the diversity of retinal conditions present in this dataset.

Demographic data including age, sex, socioeconomic status estimated using the index of multiple deprivation,^22^ and ethnicity, were collected where possible for all patients. This study was approved by the UK Health Research Authority (reference: 20/HRA/2158, approved 5 May 2020). Informed consent was waived, given that our study pertains to retrospective anonymized data.

### AI system overview

We used an AI system capable of multi-class segmentation and classification, described previously by De Fauw et al. to identify eyes with GA and exclude eyes with the ‘wet’ form of the disease known as choroidal neovascularization (CNV).^23^ While GA is traditionally diagnosed on color fundus photography and clinical assessment,^24^ in this study, the term ‘GA’ is used to refer to loss of the retinal pigment epithelium (RPE) with variable loss of the overlying photoreceptors on OCT, as in previous work.^23^

Briefly, this AI system processes three-dimensional OCT scans, initially segmenting anatomic features via a deep segmentation network. These tissue maps are then input to a deep classification network that outputs probabilities for the presence of multiple concomitant macular pathologies, including CNV, GA, and drusen (the early manifestation of AMD). A set of thresholds are then applied to transform these probabilities into binary diagnoses. Details of this are given in the **Supplementary Appendix**.

The model was developed using a retrospective dataset of Moorfields Eye Hospital OCT data collected between 1 June 2012 and 31 January 2017. There was no overlap in the data used to train the AI system and the data used in the current study. The model pipeline is illustrated in **Figure 2**.

**Figure 2.**
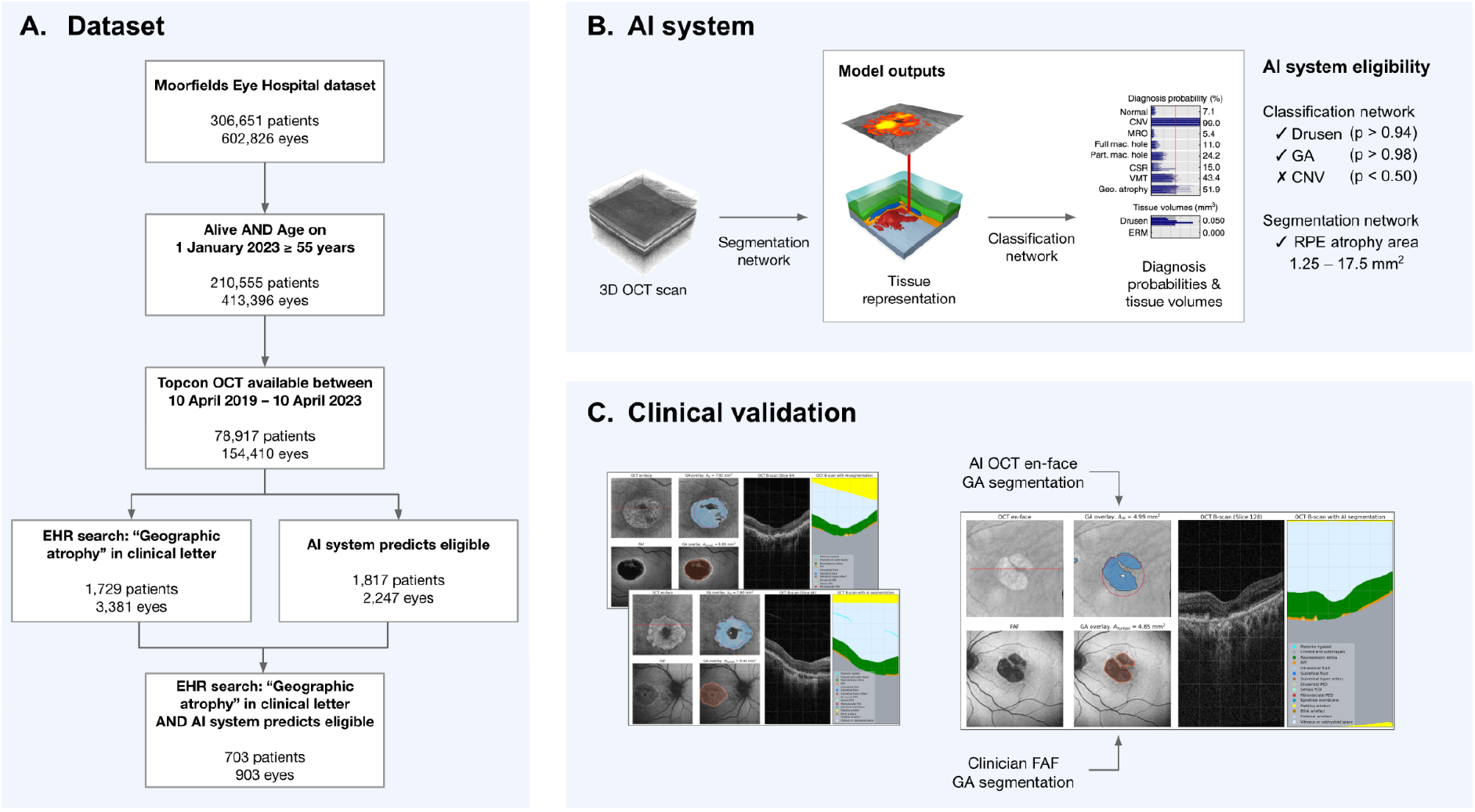
Dataset, AI system, and clinical validation overview. **A.** From a large Moorfields Eye Hospital dataset of 306,651 patients, 78,917 met our cohort inclusion criteria. The EHR search and AI system was applied to this cohort to identify a shortlist of potential participants. **B**. Whole-volume OCT scans are input to the deep segmentation network, which outputs segmentation maps of 15 anatomic and pathologic features, including the retinal pigment epithelium (RPE) layer. The deep classification network then uses these segmentation maps to predict diagnosis probabilities of 10 concomitant macular pathologies, including drusen, GA, and CNV. **C**. To estimate the positive predictive value of the shortlist of participants produced, a subset was manually graded using OCT and FAF images–the latter being manually segmented if GA was present. These were then compared to the outputs of the EHR search and OCT-based AI system.

### AI estimation of GA area

In clinical trials for GA, eligibility is typically determined by a reading center with FAF used as a gold standard to measure the area of GA.^11^ Following the first step where we confirmed the presence of GA and absence of CNV using the AI outputs, we quantified the area of GA by analyzing the AI segmentation of the RPE layer produced for each OCT B-scan. The absence of RPE was used to determine regions of GA. The total area of atrophy across the OCT was then summed across each B-scan and converted to mm^2^. This was compared to the area range specified in the trial criteria to determine if a patient should be shortlisted.

### GA location

Several trial criteria make requirements on the location of atrophy, specifically (i) whether any atrophy exists within 1500 μm from the foveal center, which we termed ‘central’ GA, and (ii) whether atrophy is found at the center point of the fovea, which we termed ‘fovea involving’ GA.

To determine these criteria we needed to locate the anatomical fovea in the OCT scan. Topcon OCT scans were fixated on the preferred retinal locus – the area of the retina which individuals naturally use to fixate on an object. In eyes with macular conditions such as GA, this may not correspond with the anatomical fovea.^25^ Instead, we approximate the foveal coordinates using the neurosensory retina (NSR) segmentation produced by the AI system. Further details of this approach are available in the **Supplementary Appendix**.

### Clinical trial criteria

To showcase our AI-based approach for shortlisting patients, we selected relevant criteria from the HORIZON trial. HORIZON was a phase II clinical study evaluating the safety and efficacy of a single subretinal injection for patients with GA.^26^ Eligibility was met if an individual was (i) aged over 55 years old and had an eye with (ii) an absence of CNV, and (iii) GA secondary to AMD with an area between 1.25–17.5 mm^2^ inclusive. These criteria were used to filter the AI outputs and produce a shortlist.

We expanded our analyses to three additional trials with distinct criteria to fully showcase the capabilities of the AI system. Two of these trials supported the recent FDA approvals of pegcetacoplan^12^ (DERBY)^27^ and avacincaptad pegol^11^ (GATHER2)^28^, with a third trial by Janssen currently underway.^29^ The criteria selected to evaluate the AI system for each trial are listed in **Table S1**.

### EHR search

A common approach to identify eligible trial participants is via a keyword search of the EHR.^30^ Patients attending Moorfields Eye Hospital receive clinical letters following each visit. We performed a keyword search of these letters to identify patients with the keyword ‘geographic atrophy’ appearing in at least one clinical letter. The earliest date on which the keyword appeared in a patient’s record was used to flag any subsequent retinal scans.

### Clinical validation

We validated the AI diagnoses and area measurements against expert evaluations based on HORIZON trial criteria by randomly selecting patients across three groups: (A1) AI-shortlisted and affirmative EHR result; (A2) AI-shortlisted and negative EHR result; (B1) affirmative EHR result but not AI-shortlisted. For a patient to be available for sampling, an OCT and FAF image, acquired within 90 days on either side of the OCT, must have been available for both eyes.

Experts graded image quality and the presence of drusen, GA, and CNV using OCT and FAF. Grading was performed by a senior grader specializing in retina with 10 years of experience (R.C.) and a medical retina fellow with 10 years of clinical experience (M.J.). Disagreements regarding diagnosis were arbitrated by a senior retina specialist with 20 years of experience (P.A.K.). Following adjudication, in good-quality images without CNV but with GA, the borders of all areas of definite decreased autofluorescence (DDAF) corresponding to GA on FAF were manually outlined using ImageJ.^31^ The total segmented GA area in mm^2^ was averaged between clinicians.

Bland-Altman plots were used to judge the agreement between the averaged FAF segmentation area and AI-based OCT segmentation, as well as between the segmentation areas of each grader. Intraclass correlation coefficients (ICC) were calculated using the two-way mixed-effects model for agreement. All 95% confidence intervals (CIs) were calculated via bootstrapping with 10,000 samples.

### Analysis and inference

Our goal was to estimate the proportion of patients who would be truly eligible for the trial, based on the imaging inclusion criteria, from all of those who were shortlisted. After inferring the positive predictive value (PPV) for the cohort of patients shortlisted, we performed a weighted sum between the appropriate values from the clinical validation and the total number of patients shortlisted. More details can be found in the **Supplementary Appendix**.

## Results

### Cohort demographics

From the dataset of 78,917 patients (154,410 eyes), 1,817 patients (2,247 eyes) and 1,729 patients (3,381 eyes) were shortlisted by the AI system and the EHR search, respectively. Patient demographics are described in **Table 1**.

**Table 1.** Patient demographics. Age data were calculated based on an individual’s age on 1 January 2023. Sex data were missing for 10 individuals in (A). Socioeconomic data were missing for 7.3%, 12.2%, and 13.0% of individuals from datasets (A), (B) and (C) respectively. *P* values were obtained via bootstrapping with 10,000 samples. All pairwise tests with (A) returned *P* values < 0.001 aside from ‘Ethnicity - Other’ which returned *P* values of 0.015 and 0.028 with (B) and (C) respectively.

| Demographic |  | (A) Dataset -<br>passed<br>inclusion<br>criteria<br>(78,917<br>patients) | (B) AI system<br>- Shortlisted<br>(1,817<br>patients) | (C) EHR<br>search -<br>Shortlisted<br>(1,729<br>patients) | P value -<br>comparing<br>(B) to (C) |
| --- | --- | --- | --- | --- | --- |
| Age | Mean (SD) | 72.7 (10.4) | 82.3 (9.2) | 84.0 (8.4) | <0.001 |
| Socioeconomic<br>status | Mean (SD) | 5.2 (1.8) | 5.6 (1.9) | 5.6 (1.9) | 0.723 |
| Sex (%) | Female (%) | 42,158 (53.2) | 1,182 (65.1) | 1,115 (64.5) | 0.261 |
| Ethnicity (%) | Unknown (%) | 24,428 (30.8) | 445 (24.5) | 364 (21.1) | 0.054 |
|  | White (%) | 22,525 (28.4) | 884 (48.7) | 895 (51.8) | 0.284 |
|  | Asian (%) | 12,769 (16.1) | 196 (10.8) | 201 (11.6) | 0.311 |
|  | Mixed (%) | 12,690 (16.0) | 5 (0.3) | 1 (0.1) | 0.618 |
|  | Black (%) | 6,424 (8.1) | 46 (2.5) | 28 (1.6) | 0.047 |
|  | Other (%) | 478 (0.6) | 241 (13.2) | 240 (13.9) | 0.464 |

### AI system vs EHR

Of the 1,729 patients shortlisted by the EHR search, inference calculations based on the clinical validation estimated that 693 (40%; 95% CI: 39–42%) from this group would have ultimately fulfilled the HORIZON imaging criteria. Of the 1,817 patients shortlisted by the AI system, 1,139 (63%; 95% CI: 54–71%) from this group would have ultimately fulfilled the trial criteria. Compared to the EHR search, our AI system achieves a 23% absolute improvement (*P* < 0.001) in the proportion of eligible patients found from those shortlisted.

A combined search using the EHR and AI system together shortlisted 703 prospective candidates. Of this group, estimates showed 604 patients (86%; 95% CI: 79–92%) would match the eligibility criteria, resulting in a 46% absolute improvement over the EHR search alone (*P* < 0.001). These results are detailed in **Table S2**.

### Extending to other clinical trials

Beyond meeting the imaging criteria for the HORIZON trial, the AI system is capable of adjusting to additional criteria across several other GA clinical trials. Differences include allowed age ranges, the extent to which study or fellow eye CNV is considered exclusionary, the allowed limits of GA area, and whether patients are eligible based on the location of GA relative to the fovea. A summary of each eligibility criteria, as well as the number of patients the AI shortlisted for each trial, can be found in **Table S1**. Compared to the 1,817 patients shortlisted for the HORIZON trial, our AI system shortlists 1,580 patients for DERBY, 768 for the Janssen trial, and 438 for GATHER2. We present examples of AI segmentations for patients who were shortlisted for each trial in **Figure 3**.

**Figure 3.**
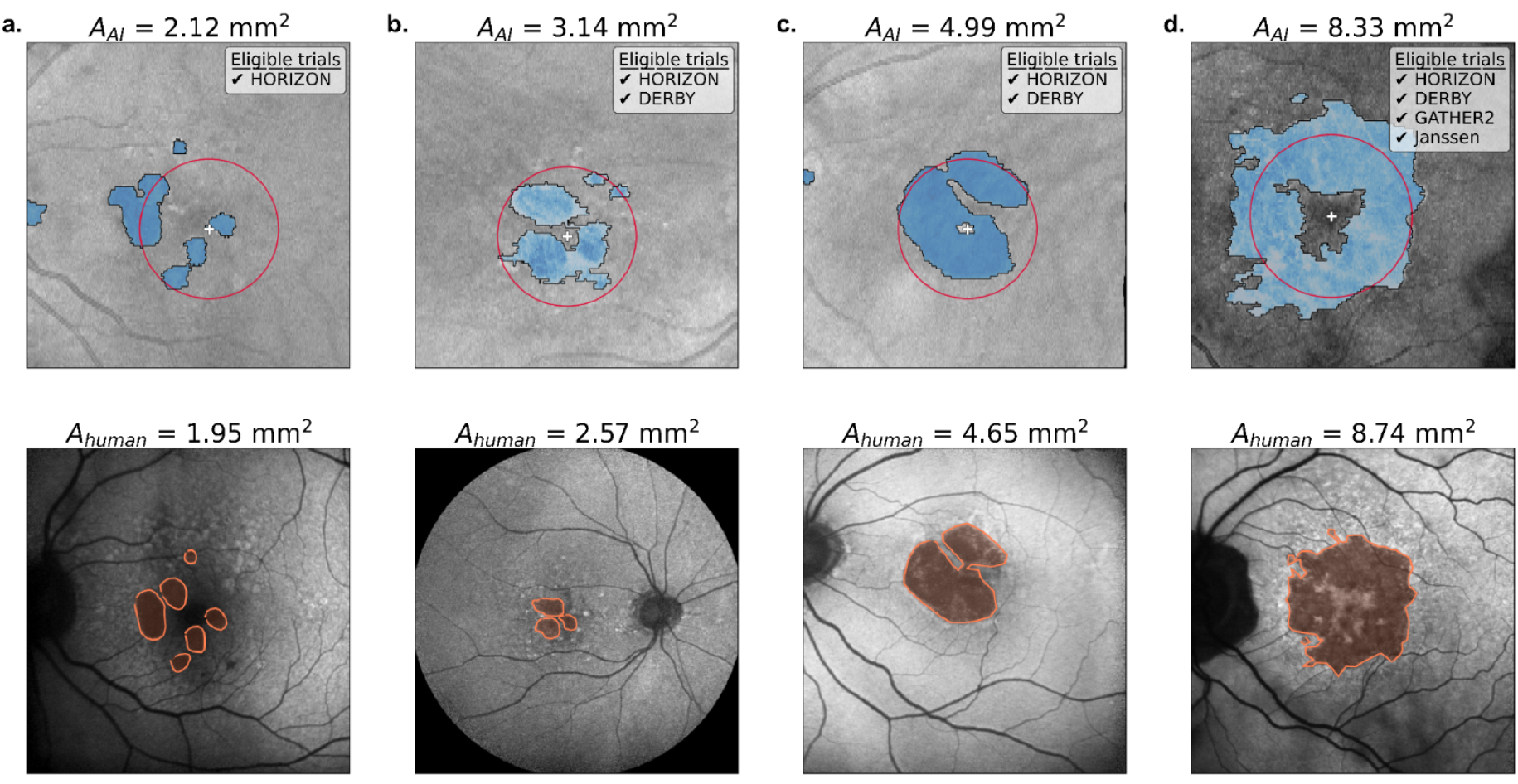
Examples of the AI-segmented OCT and clinician-segmented FAF. Each subplot contains the OCT en-face overlaid with the AI segmentation (top) and the associated FAF image from the same eye overlaid with the segmentation from grader R.C (bottom). The AI-determined fovea location (white plus) and a 1500 μm circle centered on the fovea (red circle) are indicated. The AI and averaged human GA area estimations are indicated, as well as the clinical trials each individual is eligible for based on the AI results (taking into account AI outputs for both eyes where necessary). **(a)** This patient is solely eligible for HORIZON since *A*_*AI*_ is below the lower limit for the other trials (2.5 mm^2^). Patients in **(b)** and **(c)** have central and non fovea-involving GA, but CNV in their fellow eyes excluding them from GATHER2 and the Janssen trials. The patient in **(d)** was determined by the AI to fit the imaging criteria for all trials.

### Clinical validation of the AI system

The accuracy of predicting clinical trial eligibility was evaluated using PPV and NPV values derived from the clinical validation (**Table 2**). Using human evaluation as the gold standard reference, the AI system shortlists patients with a PPV of 86% (95% CI: 79–92%) when “geographic atrophy” was present in the EHR and 48% (95% CI: 38–57%) when it was absent. Only 9% (95% CI: 3–16%) of patients with an affirmative EHR search but not shortlisted by the AI were found to be eligible. As described further in the **Supplementary Appendix**, these results were used to calculate the yields in the inference process above.

**Table 2.** Clinical validation results. Each stratum was defined by whether a patient had an affirmative EHR result at a visit prior to the included scan, and whether the AI system determines that at least one of a patient’s eyes is eligible based on the HORIZON trial criteria. Patients were split into one of these three strata based on their most recent eye scans and then sampled randomly to form the clinical validation datasets.

| Number of eligible patients | (1) EHR contains ‘geographic atrophy’ |  | (2) EHR does not contain ‘geographic atrophy’ |  |
| --- | --- | --- | --- | --- |
| (A) AI shortlisted | 84 / 98 | PPV: 86%<br>(95% CI: 79–92%) | 36 / 75 | PPV: 48%<br>(95% CI: 38–57%) |
| (B) AI did not shortlist | 7 / 81 | PPV: 9%<br>(95% CI: 3–16%) |  |  |

Agreement between AI-derived area and the averaged grader-segmented area is illustrated in **Figure S2**. Comparing GA area showed a mean difference of 0.70 mm^2^ (95% CI: -6.08– 7.47 mm^2^), with an intraclass correlation coefficient (ICC) of 0.70 (95% CI: 0.61–0.78). Restricting analysis to the eligible area range showed a mean difference of 0.27 mm^2^ (95% CI: -5.21–5.75 mm^2^), with an ICC of 0.77 (95% CI: 0.68–0.84). Inter-grader Bland-Altman plots can also be found in **Figure S2**. Comparing GA area showed a mean difference of - 0.41 mm^2^ (95% CI: -5.65–4.82 mm^2^), with an ICC of 0.92 (95% CI: 0.89–0.94). Restricting analysis to the eligible data points showed a mean difference of -0.15 mm^2^ (95% CI: -2.32– 2.02 mm^2^), with an ICC of 0.97 (95% CI: 0.95–0.98).

The median GA area of those shortlisted by the AI was 8.56 mm^2^ (95% CI: 8.22–8.96 mm^2^) (**Figure S3**). The subset with an affirmative EHR search had a median GA area of 10.1 mm^2^ (95% CI: 9.36–10.6 mm^2^), while those without the EHR search had a median area of 7.52 mm^2^ (95% CI: 6.95–8.02 mm^2^).

## Discussion

AMD remains a very common cause of poor vision in the elderly and there is currently intense research activity aiming to slow down cell death in the advanced stages of GA with novel treatments.^32, 21^ In this study, we demonstrate that AI can sift through large amounts of diverse, real world data to identify candidates meeting specific image-based selection criteria for GA trials. The AI system also accurately quantifies the GA area using OCT scans, exhibiting good agreement with expert-graded FAF images.

Conventional EHR search strategies that rely on clinical notes may overlook patients with GA on imaging but whose diagnosis is not explicitly recorded by their ophthalmologist. They may also capture patients with GA documented in their notes who subsequently fail to meet the rigorous image-based criteria for trial eligibility. In our study, this method produced an eligibility rate of 40%. Using our AI algorithm alone boosts the eligiblity rate to 63%. When combined with the EHR search, this increases further to 87%, meaning our AI-based approach significantly reduces the likelihood of an individual experiencing an imaging-based screening failure. Our approach has the potential to streamline the patient shortlisting process, offering a more efficient and precise alternative to a traditional EHR search as well as enabling clinics without sophisticated EHR systems to benefit from automated shortlisting.

From an initial cohort of almost 80,000 patients, using our combined AI-EHR approach, we identify over 600 patients with a high likelihood of imaging eligibility for the HORIZON trial. Our AI system can also adjust to meet specific image-based criteria of other GA trials, like GA location (central or foveal) and the presence of CNV. This allowed us to produce tailored shortlists of patients for the DERBY/OAKS, GATHER2, and Janssen trials.^11,12, 29^ When applied with predetermined criteria, our approach enables trial sponsors to assess site feasibility by estimating patient enrollment figures based on the resources available to screen and the disease prevalence, thereby facilitating a data-driven approach to protocol design.

The AI model estimates the size of GA by evaluating the area of RPE loss. We found that there was good agreement in the cross-modality (FAF/OCT) comparison between the AI and human graders.^33^ Despite being inferior when compared to the inter-grader agreement across the same modality, the AI-grader results achieve an ICC in the range of previous studies which examine automated segmentation of OCT scans as a surrogate for manually-graded FAF.^34^ One inherent limitation with this approach is that FAF and OCT imaging have fundamental differences which may lead the AI to over-or under-estimating the total GA area. FAF provides information on the condition of both the RPE and photoreceptors without the ability to evaluate each structure individually.^35^ When human graders measure the area of DDAF on FAF, it might not reflect the actual condition of the RPE alone. In contrast, the OCT is depth resolved and evaluates RPE loss specifically. We found that the AI tended to underestimate the total GA area for lesions larger than 17.5 mm^2^ on FAF, especially when they extended beyond the OCT field of view **(Figure S4)**. Registration between each FAF and OCT pair would help to mitigate this. Overestimations were also observed, potentially caused by errors in RPE segmentation, such as around the optic nerve due to peripapillary atrophy **(Figure S5)**.

In conclusion, we demonstrate that an AI tool can facilitate clinical trial recruitment in AMD. Given the intense clinical trial activity currently underway, we believe that implementing this AI-driven strategy will offer a scalable solution to the recruitment challenges in GA trials. Our future efforts will concentrate on assessing the real-world robustness of this AI solution, ensuring it performs well with routinely collected images of varying quality, protocols, equipment, and across diverse populations.

## Data Availability

All data produced in the present study are available upon reasonable request to the authors.

## Notes

**Financial Disclosures:** Dr. Keane has acted as a consultant for Google, DeepMind, Roche, Novartis, Apellis, and Bitfount and is an equity owner in Big Picture Medical. He has received speaker fees from Heidelberg Engineering, Topcon, Allergan, and Bayer. He is supported by a Moorfields Eye Charity Career Development Award (R190028A) and a UK Research & Innovation Future Leaders Fellowship (MR/T019050/1). Dr Wagner is funded by a Medical Research Council Clinical Research Training Fellowship (MR/T000953/1). Dr. Guymer is a Consultant to Roche, Genentech and on advisory boards for Apellis, Roche, Genentech, Bayer, Novartis, Belite Bio, Ocular Therapeutix, Complement Therapeutics, Boehringer Ingelheim Pharmaceuticals and Character Bioscience. Dr. Wu and Dr. Guymer are supported by the National Health & Medical Research Council of Australia (#2008382 [Z.W.] and #1194667 [R.G.]).

**Funding Support:** This work was supported by Innovate UK grant number 1008100. Dr. Keane is supported by UK Research & Innovation Future Leaders Fellowship (MR/T019050/1)

### Competing Interest Statement

The authors have declared no competing interest.

### Funding Statement

This work was supported by Innovate UK grant number 1008100. Dr. Keane is supported by UK Research & Innovation Future Leaders Fellowship (MR/T019050/1).

### Author Declarations

This study was approved by the UK Health Research Authority (reference: 20/HRA/2158, approved 5 May 2020). Informed consent was waived, given that our study pertains to retrospective anonymized data.

