## Supplementary Appendix for "Artificial intelligence to facilitate clinical trial recruitment in age-related macular degeneration"

### Supplementary Methods

#### Dataset

From the original cohort of 306,651 patients (602,826 eyes) who attended Moorfields Eye Hospital, we applied the following criteria for inclusion in this study: patients who were alive and aged over 55 years on 1 January 2023, and had a Topcon (Tokyo, Japan) macula-centered OCT volume scan with at least 128 B-scans using the 3D OCT-2000 or DRI OCT Triton device, between 10 April 2019 and 10 April 2023. Although not mandatory for inclusion, the dataset also contained FAF images from these patients acquired in the same period using the Heidelberg Spectralis HRA+OCT device (Heidelberg, Germany).

#### AI system operating point

AI systems which classify the presence of multiple diseases from a single image output one probability for each disease. These probabilities lie in the range 0 – 1. To convert these probabilities into binary diagnoses, disease-specific thresholds must be selected. This selection is performed using a tuning dataset, where the goal is to find the optimum threshold which delivers the best performance for a specific downstream use case. The meaning of 'optimum' here is subjective and depends on the cost associated with the AI classifying false positives versus false negatives. For example, a different set of thresholds may be more appropriate for applying the AI system in a smaller hospital, where a greater number of false positives is an acceptable trade-off in return for shortlisting additional eligible patients.

A threshold tuning set was created to determine these disease-specific thresholds. Sets of 25 OCT scans were randomly sampled for tuning based on nine strata, one for each of the three disease in the probability ranges 0.1 – 0.5, 0.5 – 0.9, and 0.9 – 1.0, totalling 225 scans. This stratified sampling helped us to more precisely determine the location of the threshold by acquiring ground truth gradings for scans near an assumed threshold. These are borderline cases where the AI is less confident in assigning to one class or the other. Stratified sampling was performed separately for each of the three classes to construct the threshold tuning set. Two clinicians single-graded the 225 OCT scans, categorizing the images based on the presence, uncertainty, or absence of drusen, GA, and CNV. Image quality of the OCT was assessed using a three-tier approach: good, borderline, and poor. The 209 images of good quality were subject to the full grading. After careful analysis of the receiver operating characteristic (**Figure S6**) and precision-recall curves (**Figure S7**), and considering the size of our dataset and the costs of misclassification, we chose a drusen threshold of 0.94, a GA threshold of 0.98, and a CNV threshold of 0.50. OCT scans with respective probabilities greater than these values were given an affirmative diagnosis by the AI.

#### AI system estimation of GA area

For eyes that pass the probability thresholds for GA, drusen, and CNV, we use the AI system to estimate the area of the GA lesion on the OCT. To achieve this we evaluated the disruption in the retinal pigmented epithelium (RPE) segmentation produced by the AI.

As these segmentations are produced for each OCT B-scan, we can create an en-face projection for the RPE segmentation specifically. Instead of plotting this as a thickness map for the RPE layer, we instead create a binary mask of the locations on the en-face where

RPE is present in the segmentation map and where it is not present. Each en-face projection has dimensions 512x512, with scans from the 3D OCT-2000 and DRI OCT Triton devices covering a 6x6 $mm^2$ and 7x7 $mm^2$ region respectively.

We consider the complete absence of RPE on the segmentation map as an indication of the presence of GA. We decided on this condition by separately comparing the areas produced when increasing the RPE thickness threshold from zero (complete absence). We produced a tuning set where a clinician was asked to grade OCT scans and, in those where GA was present, to also segment an accompanying FAF image from the same eye taken within 90 days of the OCT scan. In total, 125 OCT scan and FAF pairs were segmented. We then optimize the RPE thickness threshold to best estimate the clinician segmentations with our AI system. We found that considering only the absence of RPE to contribute to the area of GA minimized the difference to the clinician-segmented ground truth, with a median error of 1.09 $mm^2$ (95% CI: 0.80–1.49 $mm^2$), increasing as the threshold increased.

We applied basic post-processing to the binary map of atrophy to fill small gaps within larger lesions and to remove very small lesions, as we found this produced a more accurate projection. This postprocessing was restricted to regions of fewer than 264 contiguous pixels.

### GA location

As with other segmentations created by the AI system, the NSR segmentations are produced for each B-scan individually. We create an en-face projection of these NSR segmentations and approximate the location of the fovea by finding the point of minimum thickness in this projection. We validated this approach with a set of 90 OCT scans, taken from the first clinical validation strata. The coordinates of the central foveal pit (umbo) were determined by R.C. through analysis of the B-scans and used as the reference standard.

Compared to assuming the fovea is located in the image center, which had a median error to the reference standard of 0.56 mm (95% CI: 0.44–0.68 mm), our approach reduced the median error by 75% to 0.14 mm (95% CI: 0.12–0.18 mm). **Figure S8** presents illustrative examples showcasing the effectiveness of this approach.

### Clinical validation

To validate the diagnoses and area measurements provided by the AI system, we compared them to human expert grading based on the HORIZON trial criteria. We sampled 100 patients for validation based on each of the three strata A1, A2, and B1 detailed in the main text.

An eye was excluded if (1) the OCT was ungradeable, or (2) if the eye has GA and the FAF was ungradeable. A patient was excluded if both eyes were excluded, or if one eye was excluded and both eyes were required to determine eligibility. As the EHR search for ‘geographic atrophy’ is only available on a patient level, OCT and FAF images from both eyes were graded where necessary to determine a patient’s trial eligibility.

Following the assessment of OCT and FAF image quality, 98, 75, and 81 patients remain in each stratum respectively. A total of 58 patients (13, 24, and 21 from each stratum respectively) required arbitration. Reasons for arbitration were due to disagreements between graders on whether an OCT was gradable, whether a FAF was gradable, the presence of CNV, or the presence of GA.

Following analysis of the clinical validation results, strata A1 was found to have 14 disagreements between the AI and graders, A2 had 39 disagreements, and B1 had 7 disagreements. Factors contributing to these disagreements included: difference regarding the presence of CNV (43%, 31%, 14% of all disagreements in strata A1, A2, and B1 respectively); difference regarding the presence of GA (14%, 38%, 71%); and instances where the GA area fell outside of the predefined inclusion range (43%, 31%, 14%).

To produce the Bland-Altman plots in **Figure S2**, we randomly sampled and plotted the segmentation results from one eye per patient of the clinical validation. For the AI-grader plot, an eye could only be sampled if it had been segmented by the AI and segmented by the graders (i.e. the AI and graders both determined that GA was present and CNV was not present). For the inter-grader plot, an eye could only be sampled if it had been segmented by the graders (i.e. the graders determined that GA was present and CNV was not present). This resulted in 138 and 161 data points in the AI-grader and inter-grader plots respectively.

**Analysis and inference**

The clinical validation enabled estimation of the PPV and NPV values associated with each validation strata. These metrics summarize the likelihood that a patient shortlisted by the AI system will ultimately be eligible for the HORIZON trial.

Following this validation, we can calculate the number of eligible patients found by different shortlisting strategies and assess the relative efficacy of each. We choose to evaluate the three following shortlisting strategies, where:

(i) patients are shortlisted via the EHR search for ‘geographic atrophy’ alone;

(ii) patients are shortlisted via the results of the AI system alone;

(iii) patients are initially shortlisted following an affirmative EHR search, and then further analyzed by the AI system.

To estimate these values, we perform a weighted sum between the number of patients from the entire cohort that were shortlisted by the strategy, presented in **Table S3**, and the associated PPV or NPV values, presented in **Table 2**.

For each strategy, we calculate the number of eligible patients as follows:

(i) $\lfloor(703 \times 0.86) + (1026 \times (1-0.91))\rfloor$ = 696 eligible patients from a shortlist of 1729.

(ii) $\lfloor(703 \times 0.86) + (1114 \times (0.48))\rfloor$ = 1,139 eligible patients from a shortlist of 1817.

(iii) $\lfloor(703 \times 0.86)\rfloor$ = 604 eligible patients from a shortlist of 703.

These results are summarized in **Table S2**.

### Supplementary Tables and Figures

| | | Clinical trials | | | |
|---|---|---|---|---|---|
| | | **HORIZON**[26] (Novartis, Gyroscope) | **DERBY**[27] (Apellis) | **JNJ-81201887**[29] (Janssen) | **GATHER2**[28] (IVERIC bio) |
| **Eligibility criteria** | **Age** | ≥ 55 years | ≥ 60 years | ≥ 60 years | ≥ 50 years |
| | **CNV** | Not permitted in study eye | Not permitted in study eye | N/A | Not permitted in either eye |
| | **GA:** secondary to AMD | Required | Required | Required | Required |
| | **GA:** area ($mm^2$) | 1.25–17.5 | 2.50–17.5 | 2.50–17.5 | 2.50–17.5 |
| | **GA:** central | N/A | N/A | N/A | Required[36] |
| | **GA:** fovea involving | N/A | N/A | Not permitted | Not permitted |
| **Number of patients shortlisted by AI** | | 1,817 | 1,580 | 768 | 438 |

**Table S1**. **Eligibility criteria to shortlist patients for each trial with the AI system.** We define ‘central’ as being where at least 10% of the pixels in a 1500 µm radius circular area centered on the fovea contain GA. We define ‘fovea involving’ as being where 100% of the pixels in a 50 µm radius circular area centered on the fovea contain GA. The number of patients shortlisted according to each criterion is also shown.

| | **Number of patients shortlisted** | **Number of patients eligible** (95% CI) | **% of patients eligible** (95% CI) |
|---|---|---|---|
| **EHR search** | 1729 | 693 (677 - 719) | 40 (39 - 42) |
| **AI** | 1817 | 1139 (978 - 1281) | 63 (54 - 71) |
| **EHR search + AI** | 703 | 604 (555 - 646) | 86 (79 - 92) |

**Table S2**. **Estimated proportions of eligible individuals of those shortlisted.** Inference of the proportion of eligible individuals out of all those shortlisted for each strategy. 95% CIs were extrapolated from the confidence intervals calculated for the clinical validation.

| **Number of patients shortlisted** | (1) **EHR contains ‘geographic atrophy’** | (2) **EHR does not contain ‘geographic atrophy’** |
|---|---|---|
| (A) **AI predicts eligible** | 703 | 1114 |
| (B) **AI predicts ineligible** | 1026 | 76471 |

**Table S3**. **Patients filtered into validation strata.** Each cell contains the number of patients from the initial cohort belonging to each validation stratum.

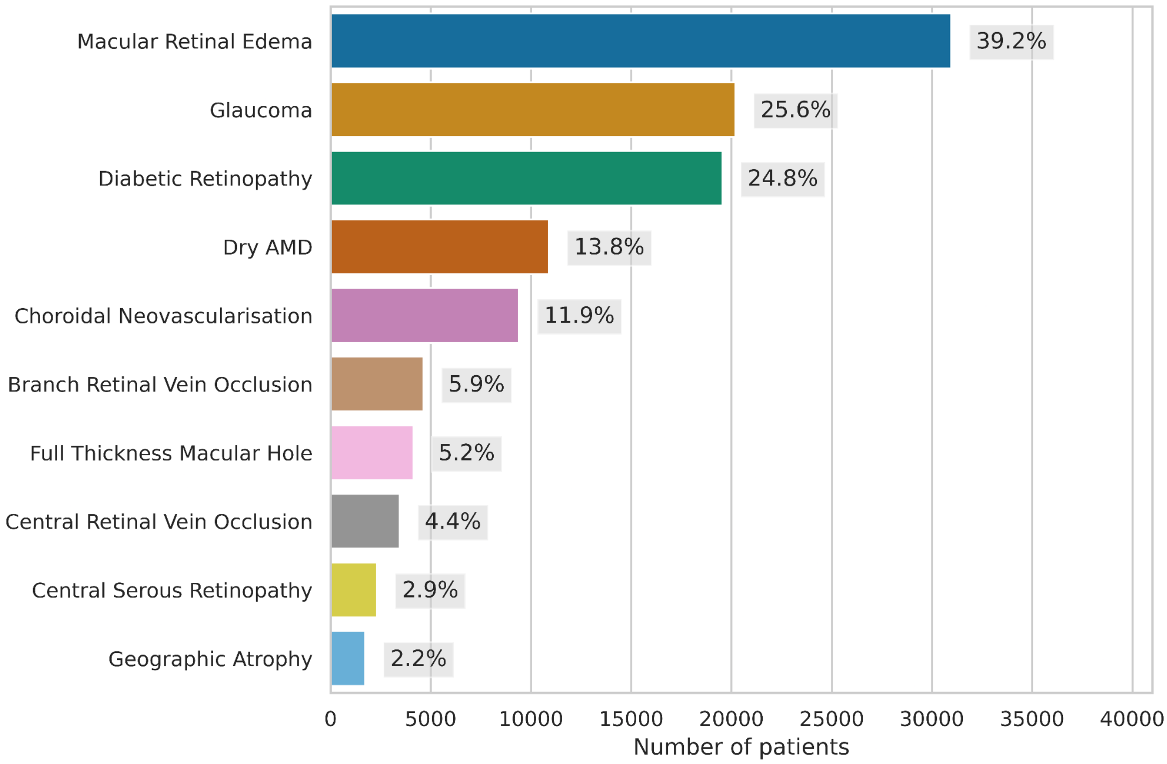


**Figure S1**. **Distribution of ten common retinal conditions as they appear in the dataset.** Each condition was identified via a keyword search in the clinical letters. Percentages were calculated based on the total number of patients passing the inclusion criteria.

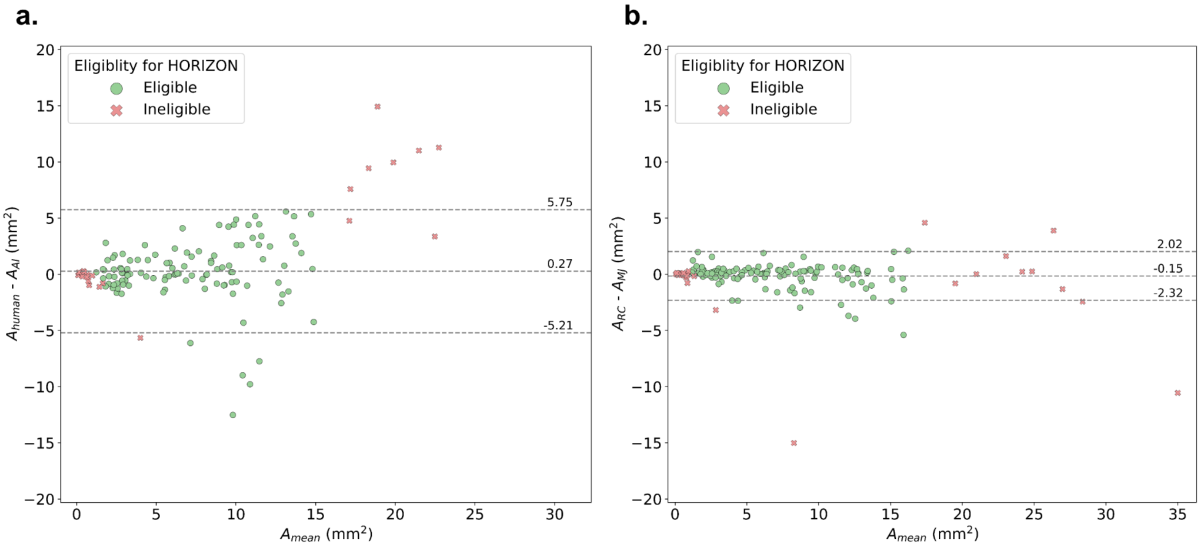


**Figure S2. Bland-Altman analysis between AI and graders. (a)** Bland-Altman plot comparing GA area computed by the AI on the OCT versus the clinician segmented area on the FAF. $A_{human}$ indicates the average measurement of both graders. **(b)** Bland-Altman plot comparing GA area segmented by graders R.C. and M.J. on the FAF. The mean difference and 95% limits of agreement are indicated, calculated based on data points eligible for HORIZON.

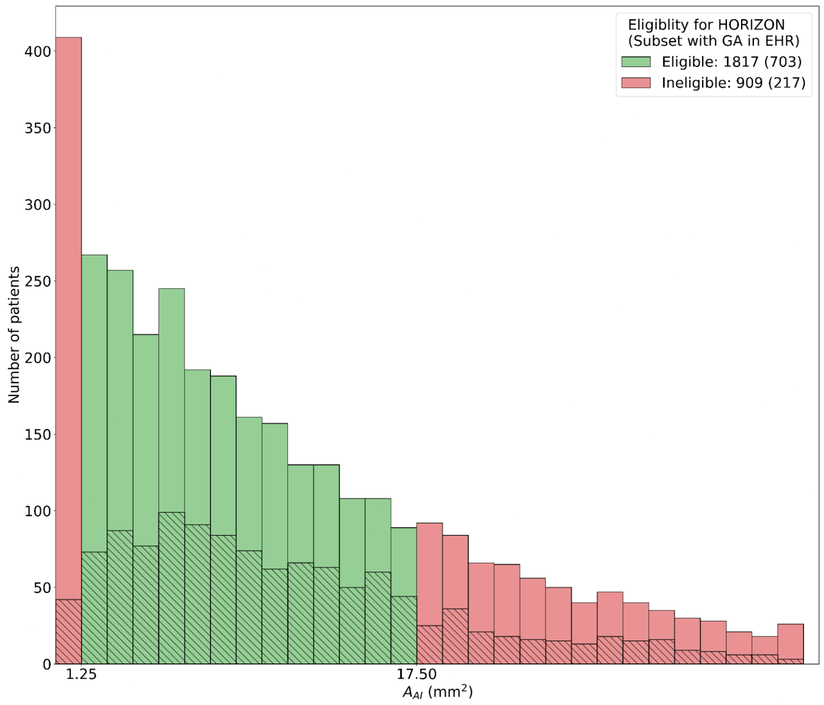


**Figure S3. Histogram of AI-segmented GA areas**. Histogram comparing the AI-segmented GA area for individuals who were determined to (i) have GA and (ii) not have CNV from the classification outputs. The hashed areas indicate the proportion of patients with an affirmative result for 'geographic atrophy' in the EHR search.

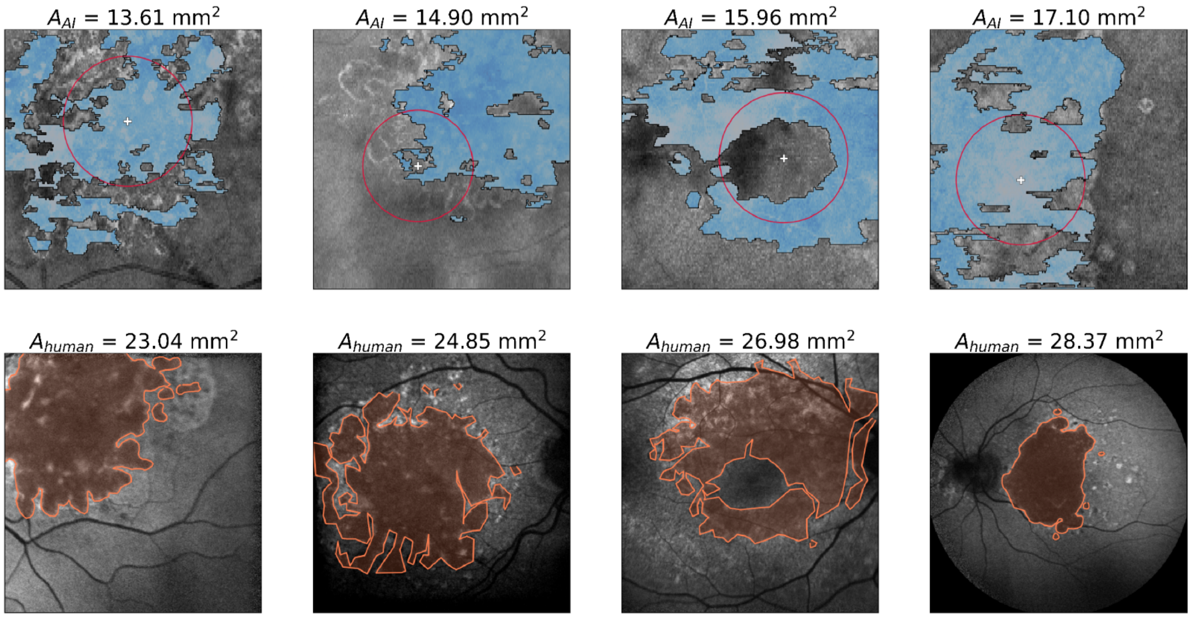


**Figure S4. Examples of the AI-segmented OCT underestimating the clinician-segmented FAF.** Shown are four examples above the upper limit of agreement in **Figure S2** (a). Images on the top row are the OCT en-face overlaid with the AI segmentation; images on the bottom row are the associated FAF image from the same eye overlaid with the segmentation from grader R.C. The AI-determined fovea location (white plus) and a 1500 μm circle centered on the fovea (red circle) are indicated. The AI and averaged human GA area estimations are indicated.

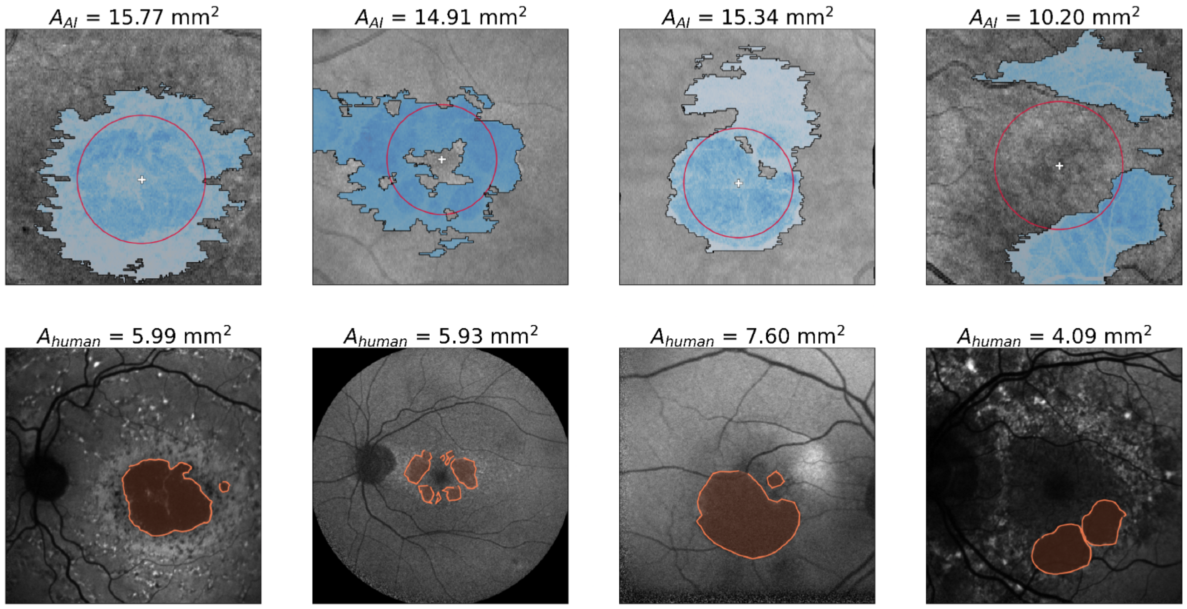


**Figure S5. Examples of the AI-segmented OCT overestimating the clinician-segmented FAF.** Shown are four examples below the lower limit of agreement in **Figure S2** (a). Images on the top row are the OCT en-face overlaid with the AI segmentation; images on the bottom row are the associated FAF image from the same eye overlaid with the segmentation from grader R.C. The AI-determined fovea location (white plus) and a 1500 µm circle centered on the fovea (red circle) are indicated. The AI and averaged human GA area estimations are indicated.

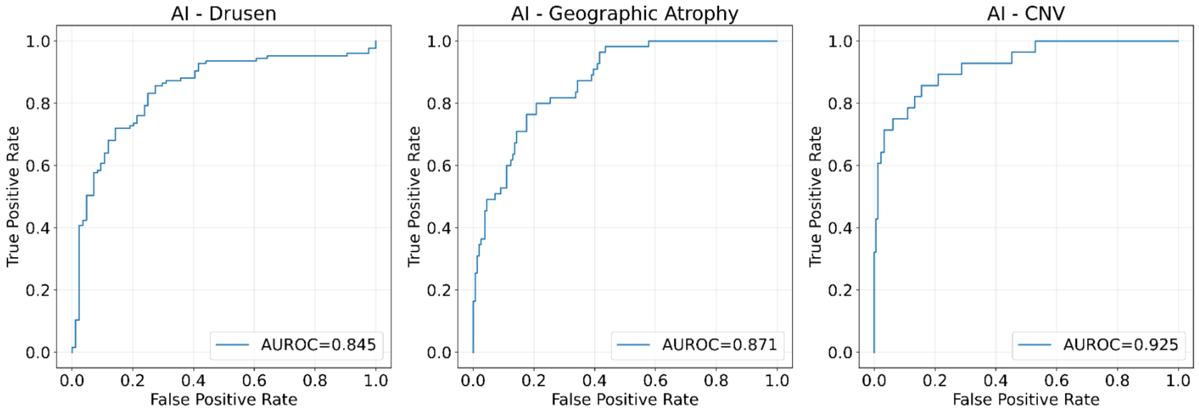


**Figure S6. Receiver operating characteristic curves from the AI system.** These were created by comparing AI predictions to the clinician-graded ground truth over the threshold tuning set.

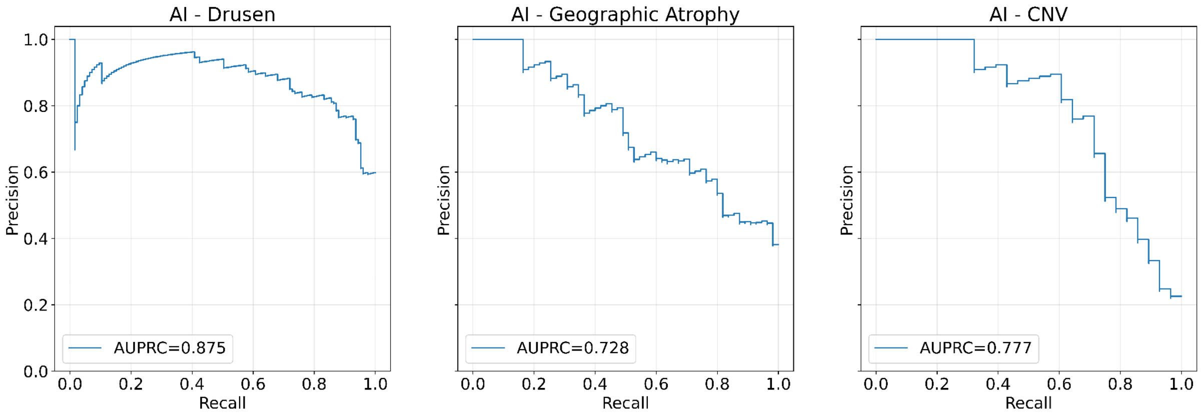


**Figure S7**. **Precision-recall curves from the AI system.** These were created by comparing AI predictions to the clinician-graded ground truth over the threshold tuning set.

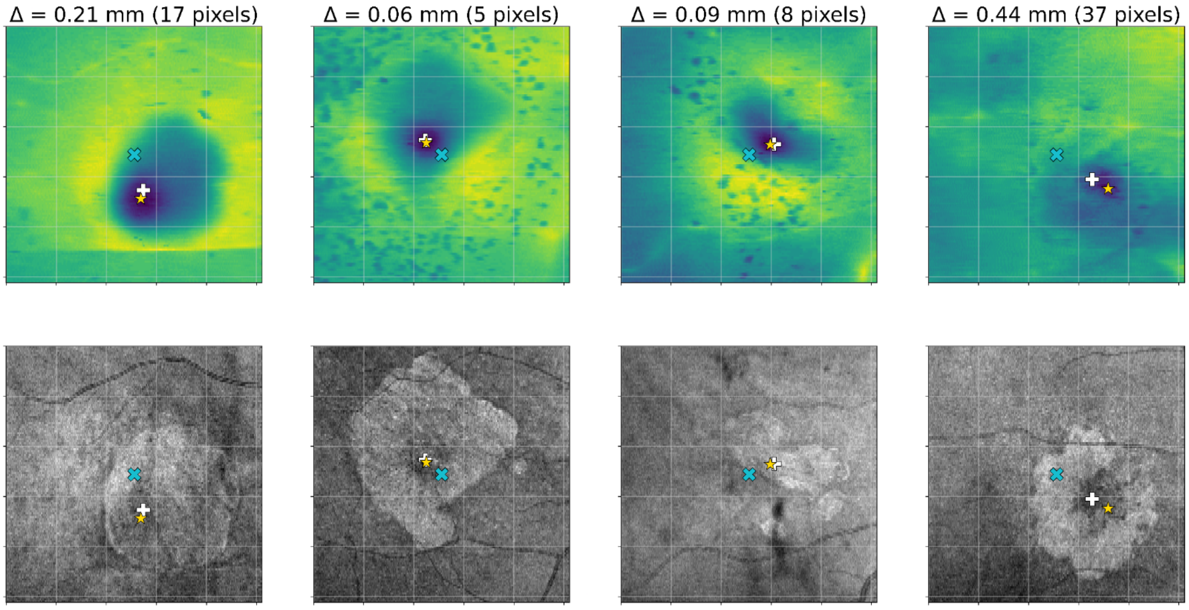


**Figure S8**. **Examples of our fovea location approximation.** Images on the top row are of the AI neurosensory segmentation map; images on the bottom row are the associated OCT en-face projection for the same scan. Comparison is between (i) assuming the fovea is located at the image center (blue cross) and (ii) the minima of the AI neurosensory retina segmentation map (white plus) with (iii) the clinician reference (gold star). The delta indicates the distance between the AI-determined location and clinician reference.
